# Combinations of multiple long-term conditions and risk of hospitalisation and death during the winter season: population-based study of 48 million people in England

**DOI:** 10.1101/2023.09.04.23295015

**Authors:** Nazrul Islam, Sharmin Shabnam, Nusrat Khan, Clare Gillies, Francesco Zaccardi, Amitava Banerjee, Vahé Nafilyan, Kamlesh Khunti, Hajira Dambha-Miller

## Abstract

**Background:** The annual winter season poses substantial challenges to the National Health Service (NHS) in England. Hospitalisation and mortality increase during winter, especially in people with multiple long-term conditions (MLTC or multimorbidity). We aimed to describe which combinations of long-term conditions (LTC) are associated with a higher risk of hospitalisation and death during winter amongst adults in England.

**Methods:** In this population-based study, we used linked primary and secondary care data from the General Practice Extraction Service Data for Pandemic Planning (GDPPR) database, Hospital Episode Statistics, and Office for National Statistics death registry. We included individuals aged ≥18 years and alive on 1^st^ December 2021 and used overdispersed Poisson models to estimate the incidence rate ratios of all-cause hospitalisations and deaths associated with the combinations of MLTCs – compared to those with no LTC – during the winter season (1 December 2021 to 31 March 2022).

**Findings:** Complete data were available for 48,253,125 adults, of which 15 million (31.2%) had MLTC. Hospitalisation per 1000 person-years was higher in individuals with MLTCs, and varied by combination, e.g.: 96, 1643, and 1552 in individuals with no LTC, cancer+chronic kidney disease (CKD)+cardiovascular disease (CVD)+type 2 diabetes mellitus, and cancer+CKD+CVD+osteoarthritis, respectively. Incidence of death (per 1000 person-years) was 345 in individuals with cancer+CKD+CVD+dementia and 1 with no LTC. CVD+dementia appeared in all the top five MLTC combinations by death and was associated with a substantially higher rate of death than many 3-, 4- and 5-disease combinations.

**Interpretation:** Risks of hospitalisation and death vary by combinations of MLTCs and are substantially higher in those with vs. without any LTCs. We have highlighted high-risk combinations for prioritisation and preventive action by policymakers to help manage the challenges imposed by winter pressures on the NHS.

**Funding:** National Institute for Health and Care Research (NIHR) through Health Data Research UK rapid funding call for the research activity “Data Science to inform NHS compound winter pressure policy response” (grant number: HDRUK2022.0313)

**Research in context:** *Evidence before this study:* We searched PubMed, from inception to April 2023, for published population-based studies examining MLTC combinations in cohorts of adults aged 18 years and over. The search terms were “multimorbidity” or ‘’multiple-long-term conditions’’ alongside “groups” or “combinations”. We found no previous studies examining MLTC in relation to death or hospitalisation during the winter season.

*Added value of this study:* We have identified distinct combinations of LTCs and estimated the associated risk of hospitalisation and deaths during the winter season using the whole-population primary and secondary care data in England.

*Implications of all the available evidence:* Understanding which combinations of MLTCs are associated with the highest risk of hospitalisation and death allows clinicians and policymakers to prioritise resources for preventative measures, such as vaccination to those that will benefit most during winter seasons.

## Introduction

Every year the National Health Service (NHS) in England faces challenges in service delivery with the onset of cold weather and an increase in acute respiratory tract infections in the winter. These challenges are usually referred to as ‘winter pressures’ and cover the period of 1^st^ December to 31^st^ March (1,2). Data from NHS England over the past decade shows a consistent pattern of high accident and emergency department attendance, prolonged wait times, and maximum hospital bed occupancy during this period (3–5). Earlier studies suggest that the demand for services and a spike in deaths during this season is, in part, being driven by an increased number of people with multiple long-term conditions (5,6).

Multimorbidity or MLTC refers to the presence of two or more long-term conditions. In 2015, an estimated 54% of people over the age of 65 years in England had MLTC and the number is projected to increase to almost 70% by 2035 (6). People with MLTC have higher service utilisation, with previous studies suggesting as much as a six-fold higher risk of attendance to accident and emergency departments compared to people with no MLTC (7–9). They are also more likely to be admitted to hospitals and then re-admitted after discharge (8,10).

Earlier large-scale studies have already established the increased service demand related to MLTC during the winter season which adds to pressures on the service (11,12). This was identified as a critical priority by the National Institute for Health and Care Research (NIHR), Health Data Research UK (HDRUK), and the Department of Health and Social Care (DHSC) (13). To date, the understanding of how MLTC affects winter pressure has been limited by the lack of granularity in the existing literature. Most studies examine the number of conditions or employ higher-level groupings of conditions rather than the granularity of individual conditions within combinations (14,15,16). Given the rising demand for services, the strain on resources, and the challenges imposed on the NHS during the winter season (17,18), understanding the impact of combinations of MLTC in relation to hospitalisation or death could highlight at-risk groups and inform prioritisation of preventive interventions. This may include, for example, vaccination strategies, for more rapid delivery to those who could benefit the most during the winter season.

In this study, we aimed to describe which combinations of long-term conditions (LTC) are associated with a higher risk of death and hospitalisation during winter amongst adults with MLTC in England.

## Methods

### Data source

In this retrospective cohort study, we used the General Practice Extraction Service (GPES) Data for Pandemic Planning and Research (GDPPR), which includes pseudo anonymised routinely collected electronic medical records from the whole population of England who are registered to a general practice. The GDPPR dataset was linked to the Hospital Episode Statistics Admitted Patient Care (HES APC) and the Office for National Statistics (ONS) death registry. These datasets are available in the Trusted Research Environment (TRE) for England established by NHS Digital (19). The TRE was accessed through the CVD-COVID-UK/COVID-IMPACT Consortium, supported by the British Heart Foundation (BHF) Data Science Centre and the Health Data Research UK (HDR UK). Further details regarding the premise of the BHF Consortium and the datasets used in this study can be found elsewhere (20).

### Study Design

Our study period was from 1 December 2021, the start of the winter pressure season, to 31 March 2022, the end of the winter season (1,2). We included all individuals registered in GDPPR who were aged 18 years or over at our study start date. Follow-up was censored at the earliest event of death or the study end date, with individuals followed up from the study start date (1 December 2021). All-cause hospital admissions (from HES APC) and deaths (from ONS deaths registry, using deaths registered by 26 January 2023) were recorded until the end of follow-up.

Within our cohort, we included only those with complete sociodemographic variables, including age at the start of the study, sex (male or female) and ethnicity (White, South Asian, Black, or Mixed/Other). Area-based socioeconomic deprivation status was denoted by the 2019 English Index of Multiple Deprivation (IMD), which is the official measure of deprivation in England (21). IMD is represented by quintiles whereby quintiles 1 and 5 represent the most and least deprived areas, respectively, and derived by mapping the 2011 Lower Layer Super Output Areas (LSOAs) from primary care records.

### Statistical analysis

As there are more than 59 different LTCs that could potentially be included under the term “multimorbidity” or “MLTC” as per recent consensus amongst UK researchers, the number of combinations (>500,000) was a challenge computationally and with limited clinical utility. We, therefore, approached the analysis in two stages:

1. We conducted data minimisation with an initial exploration, which identified more than 52,000 unique combinations (amongst 19 conditions within our cohort). To balance the computational burden with clinical utility, and taking into account the primary aim of this project, we shortlisted only the ten combinations of MLTC associated with the highest number of hospital admissions or deaths (separately) during the winter season.
2. After these ten MLTC combinations were identified, we used overdispersed Poisson regression models and estimated the incidence rate ratios with 95% confidence intervals (CI) of hospitalisation and deaths during winter associated with the combinations of MLTC, compared with those with no LTC. We used the log of the follow-up time as an offset term in the models. We estimated the crude rates of hospital admissions and deaths; to offer additional insights on the contribution of sociodemographic characteristics, we further adjusted our regression models for age, sex, ethnicity, and deprivation.

Data curation, cleaning, and exploratory analysis were performed using Python (version 3.7) and Spark SQL (version 2.4.5) on Databricks (version 6.4); statistical analysis was conducted in R (version 4.0.3).

### Ethics

This project falls within the remit of the CVD-COVID-UK/COVID-IMPACT research programme, which obtained overall ethics from the Northeast-Newcastle and North Tyneside 2 research ethics committee (REC No 20/NE/0161). Additional details of the linkage, approval, and scope of the consortium approved to use this data have been described elsewhere (22).

Patient and public contributors helped us with developing the study.

## Results

We identified a total of 50,057,280 individuals registered in GDPPR who were aged 18 years or over at the start of the study. Due to minimal missing data on sex, ethnicity, and deprivation (3.7% overall), we excluded those with missing records and conducted a complete case analysis in 48,253,125 individuals (**Figure 1**). The median follow-up was 120 days (interquartile range, IQR: 120-120). During the study period, there were a total of 4,710,675 hospital admissions and 176,895 deaths. Overall, 19.7 million individuals (40.5%) had no LTC, 13.5 million individuals (40.5%) had one LTC and more than 15 million (31.2%) people had MLTC. Individuals with MLTC were elderly (mean ± standard deviation (SD): 61.4 ± 17.9 vs. 39.9 ± 15.5 vs. 47.4 ± 16.9) compared to those with no or one LTC respectively, with a higher proportion of females (61.3% vs. 40.4% vs. 54.4%) and White ethnicity (88.7% vs. 72.5% vs. 84.0%). The distribution of deprivation was similar across the three groups (**Table 1**).

**Figure 1:**
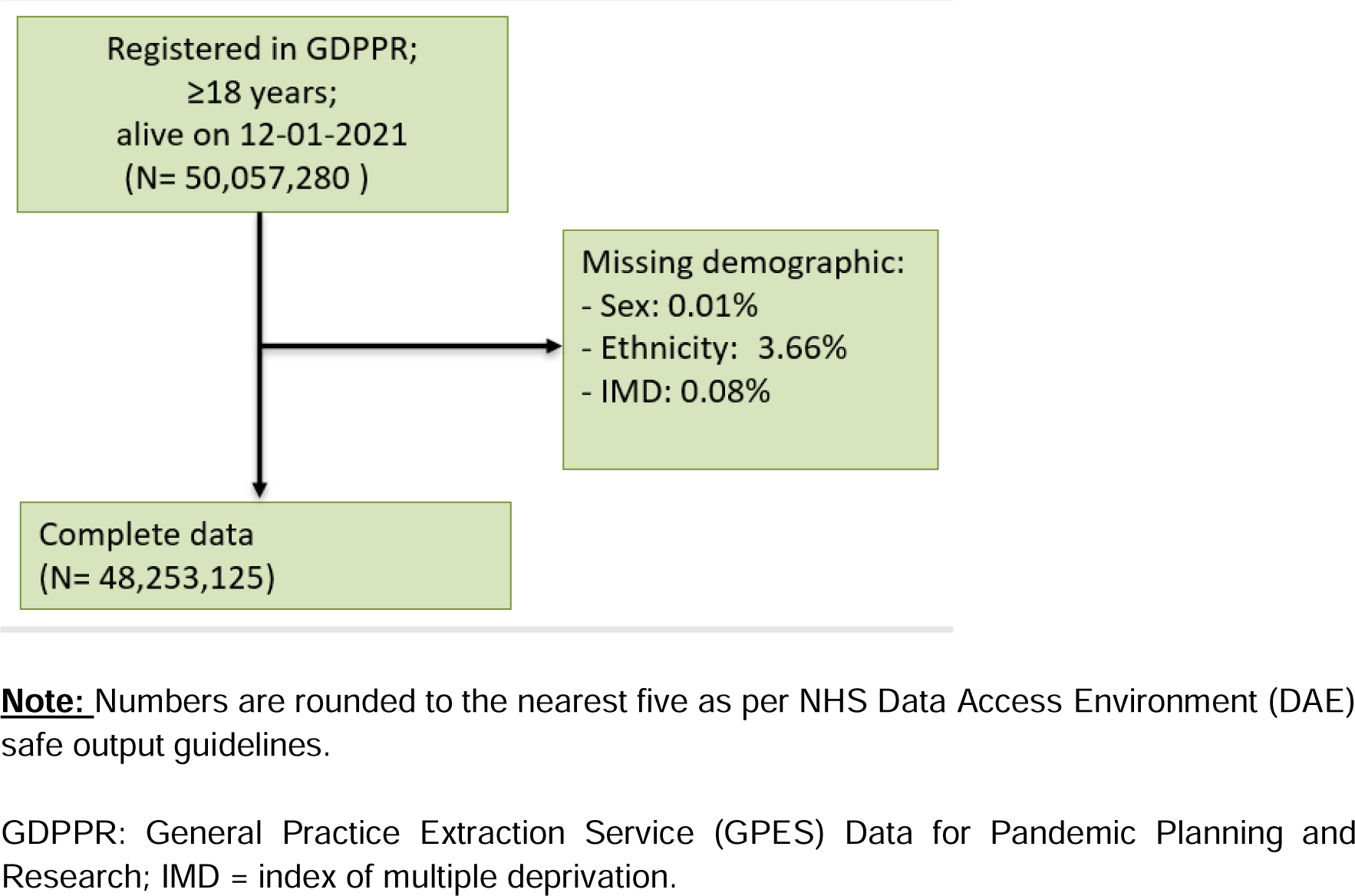
Flowchart of individuals within the cohort analysed to identify the combinations of long-term conditions associated with the highest risk of hospitalisation and deaths during winter, 2021-2022.

**Table 1:** Baseline characteristics of individuals within the cohort analysed to identify the combinations of long-term conditions associated with the highest risk of hospitalisation and deaths during winter, 2021-2022.

| **Characteristics** | **Long-term conditions** |  |  |
| --- | --- | --- | --- |
|  | **No** | **One** | **Multiple (2 or more)** |
| **N** | 19,706,155 (40.5) | 13,475,240 (27.9) | 15,071,730 (31.2) |
| **Age, mean (SD)** | 39.9 (15.5) | 47.4 (16.9) | 61.4 (17.9) |
| **Female, n (%)** | 7,962,365 (40.4) | 7,336,705 (54.4) | 9,236,450 (61.3) |
| **Ethnicity, n (%)** |  |  |  |
| Black | 1,044,605 (5.3) | 502,860 (3.7) | 426,415 (2.8) |
| Mixed/Other | 2,482,290 (12.6) | 843,050 (6.3) | 562,440 (3.7) |
| South Asian | 1,901,470 (9.6) | 806,105 (6.0) | 711,790 (4.7) |
| White | 14,277,785 (72.5) | 11,323,220 (84.0) | 13,371,085 (88.7) |
| **IMD quintile, n (%)** |  |  |  |
| 1 (most deprived) | 3,977,500 (20.2) | 2,561,345 (19.0) | 3,107,125 (20.6) |
| 2 | 4,438,705 (22.5) | 2,691,690 (20.0) | 3,033,610 (20.1) |
| 3 | 4,022,450 (20.4) | 2,749,735 (20.4) | 3,072,100 (20.4) |
| 4 | 3,716,290 (18.9) | 2,752,445 (20.4) | 3,020,925 (20.0) |
| 5 (least deprived) | 3,551,205 (18.0) | 2,720,025 (20.2) | 2,837,970 (18.8) |

**Note:** Numbers are rounded to the nearest five as per NHS Data Access Environment (DAE) safe output guidelines; SD = standard deviation; IMD = index of multiple deprivation.

### Hospital admissions

The ten combinations of MLTC that contributed to the highest rates of hospital admissions during the winter period are summarised in **Table 2**: cardiovascular diseases (CVD) appeared in only one MLTC combination; chronic kidney disease (CKD) in seven combinations; and cancer in six. We included the rate of hospital admissions for those without MLTC for comparison (**Table 2**).

**Table 2:** Number and rates of winter hospitalisation associated with the combinations of long-term conditions.

| Number of LTC | Combinations of LTC | Follow up, years | Hospital admissions, n | Incidence rate, per 1000 person-years |
| --- | --- | --- | --- | --- |
| 4 | CANCER+CKD+CVD+T2DM | 12626 | 20740 | 1643 |
| 4 | CANCER+CKD+CVD+OA | 11982 | 18595 | 1552 |
| 3 | CANCER+CKD+CVD | 29181 | 41800 | 1432 |
| 4 | AXDP+CKD+CVD+T2DM | 11434 | 13605 | 1190 |
| 2 | CANCER+CKD | 15981 | 18985 | 1188 |
| 4 | CKD+CVD+DM+OA | 14102 | 15010 | 1064 |
| 3 | CANCER+CVD+OA | 29595 | 31290 | 1057 |
| 3 | CKD+CVD+T2DM | 44354 | 39440 | 889 |
| 3 | AXDP+CKD+CVD | 21530 | 17435 | 810 |
| 3 | CANCER+CVD+T2DM | 32768 | 26495 | 809 |
| 1 | One | 4428164 | 943535 | 213 |
| 0 | None | 6477947 | 623935 | 96 |

**Note:** Numbers are rounded to the nearest five as per NHS Data Access Environment (DAE) safe output guidelines.
LTC: long-term conditions; CKD= chronic kidney disease; CVD= cardiovascular disease; T2DM=type 2 diabetes mellitus; AXDP= anxiety/depression; OA= osteoarthritis

The rate of hospitalisations (per 1000 person-years) was higher amongst individuals with MLTC compared to those without (>1600 vs 96). The highest rates of hospitalisation were found amongst the following combinations: 1643 (per 1000 person-years) for the combination of cancer+CKD+CVD+type 2 diabetes mellitus (T2DM); 1552 for the combination of cancer+CKD+CVD+osteoarthritis (OA). The rates of hospital admission increased in a dose-response manner amongst individuals who had CVD, OA, and T2DM in addition to having both cancer and CKD (**Table 2**).

Compared to those without any LTCs, the adjusted incidence rates of hospitalisation were 11.0 (95% CI: 9.4, 12.7), 9.8 (8.3, 11.4), and 9.6 (8.6, 10.7) times higher for those with cancer+CKD+CVD+T2DM, cancer+CKD+CVD+OA, and cancer+CKD+CVD, respectively. The adjusted rate of hospitalisation was 5.9 (95% CI: 5.2, 6.8) times higher in those with cancer+CVD+DM compared to those with no LTCs, which increased to 8.4 (7.0, 10.0) when anxiety/depression was further added to the MLTC combinations (**Figure 2**).

**Figure 2:**
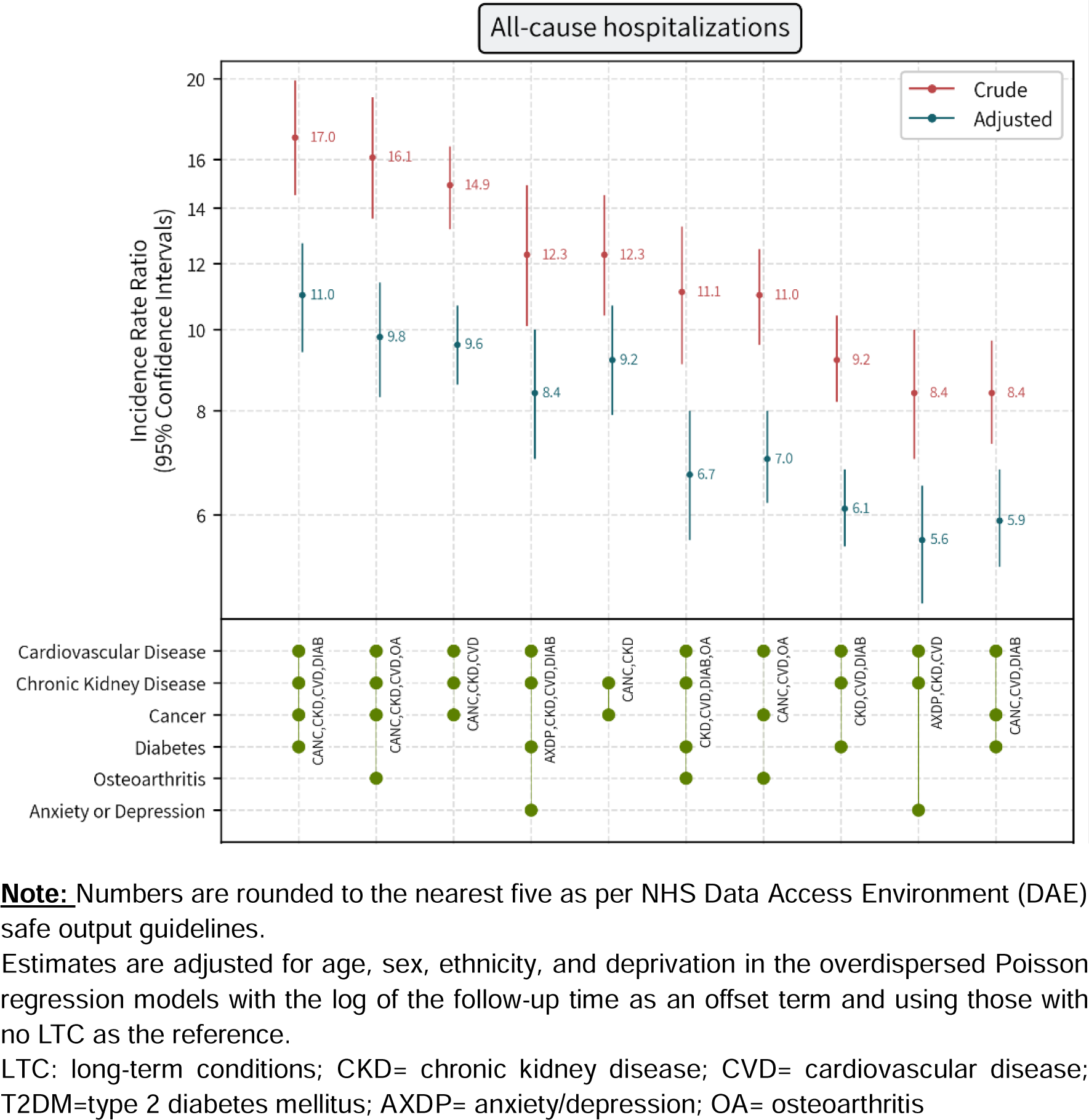
Crude and adjusted incidence rate ratios of winter hospitalisation associated with the combinations of long-term conditions compared to no long-term conditions

### Deaths

The top ten combinations of MLTC that contributed to the highest rates of mortality during the winter period are summarised in **Table 3**. CVD appeared in all MLTC combinations, while CKD appeared in seven of the top ten combinations (**Table 3**).

**Table 3:** Number and rates of winter deaths associated with the combinations of long-term conditions.

| Number of LTC | Combinations of LTC | Follow up, years | Deaths, n | Incidence rate, per 1000 person-years |
| --- | --- | --- | --- | --- |
| 4 | CANCER+CKD+CVD+DEMENTIA | 1343 | 465 | 345 |
| 4 | CKD+CVD+DEMENTIA+OA | 2101 | 700 | 334 |
| 3 | CKD+CVD+DEMENTIA | 3560 | 1030 | 289 |
| 3 | CVD+DEMENTIA+OA | 2582 | 480 | 186 |
| 2 | CVD+DEMENTIA | 6244 | 1025 | 164 |
| 5 | CANCER+CKD+CVD+DM+OA | 5717 | 690 | 121 |
| 4 | CANCER+CKD+CVD+OA | 11982 | 1310 | 109 |
| 4 | CKD+CVD+OA+OP | 5242 | 520 | 99 |
| 3 | CANCER+COPD+CVD | 5594 | 520 | 93 |
| 4 | CANCER+CKD+CVD+DM | 12626 | 1170 | 93 |
| 1 | One | 4428164 | 12970 | 3 |
| 0 | None | 6477947 | 5080 | 1 |

**Note:** Numbers are rounded to the nearest five as per NHS Data Access Environment (DAE) safe output guidelines.
LTC: long-term conditions; CKD= chronic kidney disease; CVD= cardiovascular disease; T2DM=type 2 diabetes mellitus; AXDP= anxiety/depression; OA= osteoarthritis; OP= osteoporosis; COPD: chronic obstructive pulmonary disease

The rate of deaths was 1 per 1000 person-years among people with no LTC; 345 among people with a combination of cancer+CKD+CVD+dementia; and 334 among those with a combination of CKD+CVD+dementia+osteoarthritis. CVD+dementia appeared to be the key combination that appeared in all the top five MLTC combinations. This two-disease combination was associated with a substantially higher rate of death than many 3-, 4- and 5-disease combinations. After adjusting for age, sex, ethnicity, and IMD, the rate of death was 14.6 (95% CI: 12.0, 17.8) times higher in people with CVD+dementia than in those with no LTC. The adjusted rate ratio further increased to 21.4 (95% CI: 17.5, 26.0) among patients who also had CKD (CKD+CVD+dementia), 23.2 (95% CI: 17.5, 30.3) among those who also had cancer and CKD (cancer+CKD+CVD+dementia), and 24.3 (95% CI: 19.1, 30.4) among those who additionally had CKD and OA (CKD+CVD+dementia+OA) (**Figure 3**).

**Figure 3:**
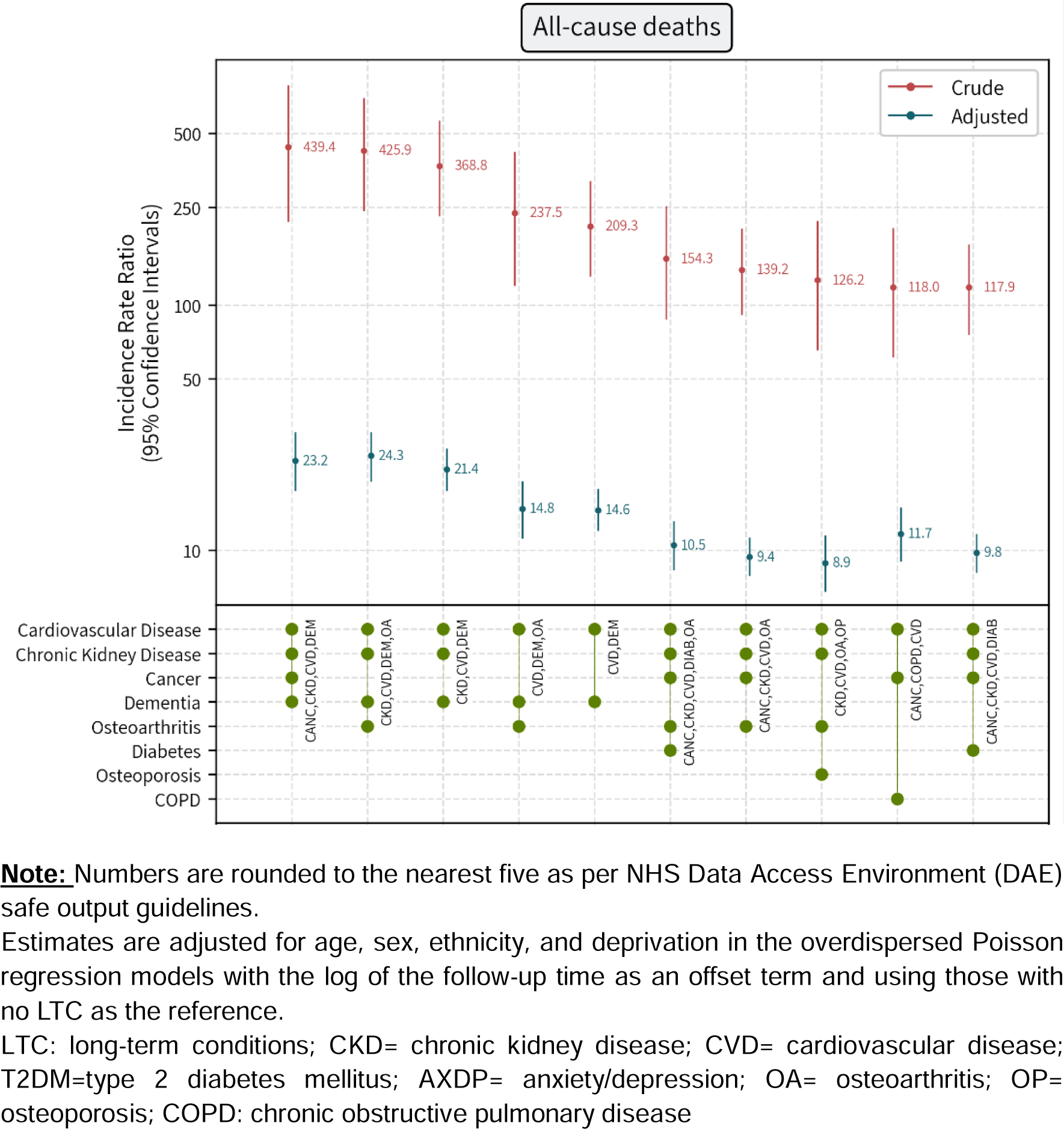
Crude and adjusted incidence rate ratios of winter deaths associated with the combinations of long-term conditions compared to no long-term conditions

## Discussion

In this large-scale whole-population study with 48,253,125 adults, the highest risk of winter hospitalisation was observed among individuals with cancer+CKD+CVD+diabetes mellitus and cancer+CKD+CVD+osteoarthritis, while the highest rate of deaths was found among those with cancer+CKD+CVD+dementia and CKD+CVD+dementia+OA.

To our knowledge, this is the largest study to date that examined how the risk of hospitalisation and deaths during the winter is associated with distinct combinations of LTC. Our study found a higher prevalence of MLTC than previously reported 27.2% (23), possibly related to the population included (the previous study used a subset of the English population while we used the whole population of England) and using different definitions of MLTC.

These results align with earlier studies that have consistently reported a significant burden of MLTC in diverse populations, with 37.2% as global prevalence and 39.2% across Europe (24–26). A recent systematic review found that in high, middle, and low-income countries there was a positive association between MLTC and hospitalization with a 2.5 times higher risk than those without MLTC (27). Our study is also the first to estimate the risks of winter hospitalisation and deaths associated with distinct combinations of MLTC. The combination that included cancer, CKD, diabetes, and CVD had the highest incidence rate ratio for the risk of hospital admissions while the combination of cancer, CKD, CVD, and dementia had the highest incidence rate ratio for the risk of all-cause deaths. The relative magnitude of the effect on both risks was slightly attenuated, but remained significantly high, after adjusting for various sociodemographic factors, including age, sex, ethnicity, and social deprivation.

The occurrence of cold weather during the winter season is associated with negative health outcomes and places significant strain on public health services. The NHS in the United Kingdom (UK) is facing growing challenges in delivering healthcare services of high quality to the population (28,29). Research conducted in the UK has revealed a pattern of increased hospital admissions during the winter months specifically for respiratory diseases, which are strongly associated with the adverse effects of cold weather and respiratory infections. Several studies conducted in the UK have also demonstrated elevated rates of hospital admissions during winter for various other conditions, including asthma, falls, specific types of road accidents, atrial fibrillation, heart failure, pulmonary embolism, stroke, and cases requiring intensive care (30–32). During the winter season, the NHS encounters capacity issues influenced by various factors beyond temperature. These factors include the rising number of patients with chronic health conditions (33,34), delays in patient transfers between different healthcare settings, and the heightened prevalence of communicable diseases like influenza, which tend to peak in winter (35–37). Hospital systems experience the most significant strain during the winter period, as hospital admissions reach their highest levels, largely due to an increase in respiratory illnesses associated with cold weather (35,38).

Our analysis identified specific combinations of chronic conditions that were strongly associated with increased hospital admissions during winter. Notably, cardiovascular diseases (CVD) were present in almost all top combinations, indicating the prominent role of CVD in driving healthcare utilization. This finding is consistent with prior research highlighting the substantial impact of CVD on hospitalizations among individuals with MLTC (39,40). Furthermore, chronic kidney disease (CKD) and cancer appeared frequently in the top combinations in our study, emphasizing their significant contributions to hospital admissions. These results corroborate existing literature that emphasizes the importance of comprehensive disease management approaches targeting CVD, CKD, and cancer to effectively reduce hospitalizations in individuals with MLTC (41,42).

A comparison of our results with previous studies further supports the robustness of these findings. However, the effect of combinations of certain diseases had not been examined before. Jani et al. conducted a study among 500,769 participants from the UK Biobank and identified cardiovascular diseases and cancer as common contributors to all-cause mortality in individuals with MLTC (42). These consistent findings across diverse populations underscore the universal burden imposed by these conditions and highlight the necessity for comprehensive management strategies addressing their co-occurrence (43–45).

In addition to hospital admissions, our study explored the association between specific MLTC combinations and all-cause death rates. The combinations of CVD with dementia and CKD demonstrated the highest rates of deaths. These findings align with previous research demonstrating the adverse impact of CVD and dementia on death (46–48). The significantly elevated death rates observed in these combinations compared to others emphasize the urgency of targeted interventions and effective management strategies for these conditions.

Comparison of our findings with existing literature further strengthens the evidence regarding the elevated death risk associated with specific MLTC combinations. Johnson et al. and Brown et al. conducted studies in different populations and reported an increased risk of death in individuals with MLTC, particularly in combinations involving CVD and dementia (49,50). These studies provide additional evidence supporting the heightened death risk associated with specific combinations of chronic conditions.

The high prevalence of MLTC and its impact on hospital admissions and death rates highlight the complex nature of managing multiple chronic conditions (46). The management of MLTC requires a comprehensive and patient-centred approach that considers the interactions between different conditions, potential polypharmacy, and the unique needs of individual patients. Integrated care models that promote collaboration between healthcare professionals and involve the active participation of patients are crucial in addressing the challenges posed by MLTC (51,52).

A person-centred approach that focuses on personalized care planning, shared decision-making, and coordinated management can help optimize outcomes for individuals with MLTC (53). Collaborative care models, such as the Chronic Care Model and the Guided Care Model, have shown promise in improving patient outcomes, reducing hospitalizations, and enhancing the quality of life for individuals with MLTC (54,55). These models emphasize care coordination, self-management support, and enhanced communication between patients and healthcare providers.

Furthermore, interventions aimed at addressing specific combinations of chronic conditions that contribute to the highest healthcare utilization and death rates are crucial. For instance, interventions targeting CVD, CKD and dementia should prioritize the management of cardiovascular risk factors, including hypertension, diabetes, and hyperlipidaemia, while also addressing cognitive impairment and promoting brain health (43–45)(39). Cancer care pathways should be tailored to address the unique needs of individuals with MLTC, considering potential interactions between cancer treatments and other chronic conditions (56,57).

Until now, the largest study on MLTC in the UK included 1.1 million primary care patients in England (58). The study concluded that the number and selection of conditions resulted in very large differences in MLTC prevalence which, was 35.2% considering the 20 most common conditions (58). Our large-scale study uses the highest-ever sample size to report MLTC status in the UK and provides compelling evidence regarding the substantial burden of MLTC on hospital admissions and death rates during winter. The identification of specific combinations of chronic conditions associated with increased healthcare utilisation and death offers valuable insights for healthcare professionals and policymakers in developing targeted interventions and strategies to reduce hospital burden during the winter season. Future research should focus on longitudinal studies to elucidate the temporal patterns and long-term impact of MLTC on health outcomes. Furthermore, efforts should be directed towards developing integrated care models that address the complex needs of individuals with MLTC during winter, particularly those with high-risk combinations of chronic conditions.

### Strengths and limitations

A key strength of our study is the large sample size of 48 million people covering most of the population in England. Our sample is representative, generalisable, and rapidly applicable to winter pressure planning over the coming year. To date, this is the largest study about MLTC in England with few missing data. We have also defined the LTCs using a combination of primary and secondary care data. Our study has several limitations, including dependence on the electronic health records for coding of MLTC: these are routine clinical records and not necessarily of research standard; furthermore, there was no grading on conditions, indicating possible over or under-representation of conditions and their severity. Conditions managed through self-care, over-the-counter treatment, private clinics, or screening programmes may have not been captured in these records. Second, our study focused on selecting LTCs to inform public health policy. The inclusion of additional LTC, especially rare diseases, would increase the overall prevalence of MLTC. Third, our study did not consider the duration or severity of illness, nor did it address the sequence of LTC.

## Conclusions

MLTC is associated with a higher risk of hospitalisation and death yet this risk varies across different conditions. Current policy and clinical guidance consider the MLTC risk of hospitalisation and death during the winter season as a single, homogenous condition. By highlighting specific high-risk combinations, our findings will inform winter pressures planning and appropriate resources (e.g., preventing screening, vaccination) where they are needed the most.

## Data Availability

The analytical codes and phenotypes used within the NHS Digital Trusted Research Environment are available in the following repository (GitHub link https://github.com/BHFDSC/CCU059_01). This project used anonymised electronic health records and administrative data which were collected and curated by NHS Digital in a Trusted Research Environment. Due to policies on information governance restrictions, the authors are unable to share individual patient data directly, but data access is available for research conditional on the approval of a research proposal and protocol, data access agreements with NHS Digital, and other information governance requirements. The authors and colleagues across the CVD-COVID-UK/COVID-IMPACT consortium have invested considerable time and energy in developing this data resource and would like to ensure that it is used widely to maximise its value. For inquiries about data access, please see https://web.www.healthdatagateway.org/dataset/7e5f0247-f033-4f98-aed3-3d7422b9dc6d. Data access approval was granted to the CVD-COVID-UK consortium (under project proposal CCU059) through the NHS Digital online Data Access Request Service (DARS-NIC-391419-J3W9T). NHS Digital data have been made available for research under the Control of Patient Information notice, which mandated the sharing of national electronic health records for COVID-19 research (https://digital.nhs.uk/coronavirus/coronavirus-covid-19-response-information-governance-hub/control-of-patient-informationcopi-notice)

## Acknowledgement

We would like to thank our public contributors for their contributions. We would also like to thank our policy colleagues from the HDRUK (Rouven Priedon and Mehrdad Mizani) and DHSC (Ruadhan Parnell and Oram Lottie) for their help and critical feedback on the analytic approach and findings that are most relevant, pragmatic, and actionable at the whole population level. KK, CLG, FZ, and SS are supported by the National Institute for Health Research (NIHR) Applied Research Collaboration East Midlands (ARC EM), NIHR Global Research Centre for Multiple Long-Term Conditions and the NIHR Leicester Biomedical Research Centre (BRC).

## Author contributions

NI and HDM conceived the project, designed the study, interpreted the outputs and wrote the original draft. SS carried out the data curation, cleaning, exploratory data analysis, and data visualization. NK contributed towards the initial draft of the paper. NI carried out the analysis and produced all outputs including detailed aggregated data and statistical outputs. All authors reviewed analysis outputs and critically revised the manuscript.

## Data and code sharing

The analytical codes and phenotypes used within the NHS Digital Trusted Research Environment are available in the following repository (**GitHub link** https://github.com/BHFDSC/CCU059_01). This project used anonymised electronic health records and administrative data which were collected and curated by NHS Digital in a Trusted Research Environment. Due to policies on information governance restrictions, the authors are unable to share individual patient data directly, but data access is available for research conditional on the approval of a research proposal and protocol, data access agreements with NHS Digital, and other information governance requirements. The authors and colleagues across the CVD-COVID-UK/COVID-IMPACT consortium have invested considerable time and energy in developing this data resource and would like to ensure that it is used widely to maximise its value. For inquiries about data access, please see https://web.www.healthdatagateway.org/dataset/7e5f0247-f033-4f98-aed3-3d7422b9dc6d. Data access approval was granted to the CVD-COVID-UK consortium (under project proposal CCU059) through the NHS Digital online Data Access Request Service (DARS-NIC-391419-J3W9T). NHS Digital data have been made available for research under the Control of Patient Information notice, which mandated the sharing of national electronic health records for COVID-19 research (https://digital.nhs.uk/coronavirus/coronavirus-covid-19-response-information-governance-hub/control-of-patient-informationcopi-notice)

## List of abbreviations

BHF: British Heart Foundation
CKD: Chronic Kidney Disease
CVD: Cardiovascular Diseases
DHSC: Department of Health and Social Care
GDPPR: General Practice Extraction Service Data for pandemic planning
GPES: General Practice Extraction Service
HDRUK: Health Data Research United Kingdom
HES APC: Hospital Episode Statistics Admitted Patient Care
IMD: Index of Multiple Deprivation
LTC: Long term conditions
NHS: National Health Service
NIHR: National Institute for Health and Care Research
OA: Osteoarthritis
ONS: Office for National Statistics
PPIE: Public and Patient Involvement and Engagement
T2DM: Type 2 Diabetes Mellitus
TRE: Trusted Research Environment

## References

1. Charlton-Perez AJ, Aldridge RW, Grams CM, Lee R. Winter pressures on the UK health system dominated by the Greenland Blocking weather regime. Weather Clim Extrem. 2019 Sep 1;25:100218.

2. NHS winter pressures | The King’s Fund [Internet]. [cited 2023 May 25]. Available from: https://www.kingsfund.org.uk/projects/nhs-winter-pressures

3. Barnett K, Mercer SW, Norbury M, Watt G, Wyke S, Guthrie B. Epidemiology of multimorbidity and implications for health care, research, and medical education: A cross-sectional study. Lancet. 2012;380(9836):37–43.

4. Hewitt J, McCormack C, Tay HS, Greig M, Law J, Tay A, et al. Prevalence of multimorbidity and its association with outcomes in older emergency general surgical patients: an observational study. BMJ Open [Internet]. 2016 [cited 2023 May 25];6(3). Available from: /pmc/articles/PMC4823401/

5. Smith SM, Wallace E, O’Dowd T, Fortin M. Interventions for improving outcomes in patients with multimorbidity in primary care and community settings. Cochrane Database Syst Rev [Internet]. 2021 Jan 15 [cited 2023 May 25];2021(1):6560. Available from: /pmc/articles/PMC8092473/

6. Humphreys J, Jameson K, Cooper C, Dennison E. Early-life predictors of future multi-morbidity: results from the Hertfordshire Cohort. Age Ageing [Internet]. 2018 May 1 [cited 2023 May 25];47(3):474–8. Available from: https://academic.oup.com/ageing/article/47/3/474/4847357

7. Whitty CJM, MacEwen C, Goddard A, Alderson D, Marshall M, Calderwood C, et al. Rising to the challenge of multimorbidity. BMJ. 2020;368.

8. Stokes J, Guthrie B, Mercer SW, Rice N, Sutton M. Multimorbidity combinations, costs of hospital care and potentially preventable emergency admissions in England: A cohort study. PLOS Med [Internet]. 2021 Jan 13 [cited 2023 May 25];18(1):e1003514. Available from: https://journals.plos.org/plosmedicine/article?id=10.1371/journal.pmed.1003514

9. Hull SA, Homer K, Boomla K, Robson J, Ashworth M. Population and patient factors affecting emergency department attendance in London: retrospective cohort analysis of linked primary and secondary care records. Br J Gen Pract [Internet]. 2018 Mar 1 [cited 2023 May 25];68(668):e157. Available from: /pmc/articles/PMC5819981/

10. Nguyen H, Manolova G, Daskalopoulou C, Vitoratou S, Prince M, Prina AM. Prevalence of multimorbidity in community settings: A systematic review and meta-analysis of observational studies. J comorbidity [Internet]. 2019 Jan 1 [cited 2023 May 25];9:2235042X19870934. Available from: http://www.ncbi.nlm.nih.gov/pubmed/31489279

11. Millwood S, Tomlinson P, Hopwood J. Evaluation of winter pressures on general practice in Manchester: a cross-sectional analysis of nine GP practices. BJGP Open [Internet]. 2021 Jan 1 [cited 2023 May 25];5(1):1–9. Available from: https://bjgpopen.org/content/5/1/bjgpopen20x101138

12. Fares A. Winter Cardiovascular Diseases Phenomenon. N Am J Med Sci [Internet]. 2013 Apr [cited 2023 May 25];5(4):266. Available from: /pmc/articles/PMC3662093/

13. Mahase E. NHS England announces “data driven war rooms” to tackle winter pressures. BMJ [Internet]. 2022 Oct 19 [cited 2023 May 25];379. Available from: https://www.bmj.com/content/379/bmj.o2515

14. MacRae C, McMinn M, Mercer SW, Henderson D, McAllister DA, Ho I, et al. The impact of varying the number and selection of conditions on estimated multimorbidity prevalence: A cross-sectional study using a large, primary care population dataset. PLOS Med [Internet]. 2023 Apr 1 [cited 2023 May 25];20(4):e1004208. Available from: https://journals.plos.org/plosmedicine/article?id=10.1371/journal.pmed.1004208

15. Koné Pefoyo AJ, Bronskill SE, Gruneir A, Calzavara A, Thavorn K, Petrosyan Y, et al. The increasing burden and complexity of multimorbidity. BMC Public Health. 2015 Dec 12;15(1):1–11.

16. Stokes J, Guthrie B, Mercer SW, Rice N, Sutton M. Multimorbidity combinations, costs of hospital care and potentially preventable emergency admissions in England: A cohort study. PLoS Med [Internet]. 2021 Jan 13 [cited 2023 May 25];18(1). Available from: https://pubmed.ncbi.nlm.nih.gov/33439870/

17. New discharge funding and NHS winter pressures - GOV.UK [Internet]. [cited 2023 May 25]. Available from: https://www.gov.uk/government/speeches/oral-statement-on-new-discharge-funding-and-nhs-winter-pressures

18. Winter pressures guidance | Health Education England [Internet]. [cited 2023 May 25]. Available from: https://www.hee.nhs.uk/our-work/winter-pressures-guidance

19. Trusted Research Environment service for England - NHS Digital [Internet]. [cited 2023 May 25]. Available from: https://digital.nhs.uk/coronavirus/coronavirus-data-services-updates/trusted-research-environment-service-for-england

20. CVD-COVID-UK / COVID-IMPACT - HDR UK [Internet]. [cited 2023 May 25]. Available from: https://www.hdruk.ac.uk/projects/cvd-covid-uk-project/

21. The English Indices of Deprivation 2019. 2019 [cited 2023 May 26]; Available from: https://www.gov.uk/government/publications/english-indices-of-deprivation-2019-technical-report

22. Wood A, Denholm R, Hollings S, Cooper J, Ip S, Walker V, et al. Linked electronic health records for research on a nationwide cohort of more than 54 million people in England: data resource. BMJ [Internet]. 2021 Apr 7 [cited 2023 May 26];373. Available from: https://www.bmj.com/content/373/bmj.n826

23. Cassell A, Edwards D, Harshfield A, Rhodes K, Brimicombe J, Payne R, et al. The epidemiology of multimorbidity in primary care: a retrospective cohort study. Br J Gen Pract [Internet]. 2018 Apr 1 [cited 2023 May 25];68(669):e245–51. Available from: https://bjgp.org/content/68/669/e245

24. Thanakiattiwibun C, Siriussawakul A, Virotjarumart T, Maneeon S, Tantai N, Srinonprasert V, et al. Multimorbidity, healthcare utilization, and quality of life for older patients undergoing surgery: A prospective study. Medicine (Baltimore) [Internet]. 2023 Mar 3 [cited 2023 May 25];102(13):e33389. Available from: /pmc/articles/PMC10063272/

25. Chowdhury SR, Chandra Das D, Sunna TC, Beyene J, Hossain A. Global and regional prevalence of multimorbidity in the adult population in community settings: a systematic review and meta-analysis. eClinicalMedicine [Internet]. 2023 Mar 1 [cited 2023 May 25];57:101860. Available from: http://www.thelancet.com/article/S2589537023000378/fulltext

26. Kudesia P, Salimarouny B, Stanley M, Fortin M, Stewart M, Terry A, et al. The incidence of multimorbidity and patterns in accumulation of chronic conditions: A systematic review. https://doi.org/101177/26335565211032880 [Internet]. 2021 Jul 15 [cited 2023 May 25];11:263355652110328. Available from: https://journals.sagepub.com/doi/full/10.1177/26335565211032880

27. Rodrigues LP, De Oliveira Rezende ACDST, Delpino FM, Mendonça CR, Noll M, Nunes BP, et al. Association between multimorbidity and hospitalization in older adults: systematic review and meta-analysis. Age Ageing [Internet]. 2022 Jul 1 [cited 2023 May 25];51(7). Available from: https://pubmed.ncbi.nlm.nih.gov/35871422/

28. Ruane S. Integrated care systems in the English NHS: A critical view. Arch Dis Child. 2019 Nov 1;104(11):1024–6.

29. Iacobucci G. NHS in 2017:Keeping pace with society. BMJ [Internet]. 2017 [cited 2023 May 25];356. Available from: https://pubmed.ncbi.nlm.nih.gov/28057618/

30. Patterson S. Do hospital admission rates increase in colder winters? A decadal analysis from an eastern county in England. J Public Health (Bangkok) [Internet]. 2018 Jun 1 [cited 2023 May 25];40(2):221–8. Available from: https://academic.oup.com/jpubhealth/article/40/2/221/3924915

31. Levin KA, Crighton EM. Reshaping Care for Older People: Trends in emergency admissions to hospital during a period of simultaneous interventions in Glasgow City, April 2011–March 2015. Maturitas. 2016 Dec 1;94:92–7.

32. McAllister DA, Morling JR, Fischbacher CM, MacNee W, Wild SH. Socioeconomic deprivation increases the effect of winter on admissions to hospital with COPD: retrospective analysis of 10 years of national hospitalisation data. Prim Care Respir J [Internet]. 2013 Sep [cited 2023 May 25];22(3):296–9. Available from: https://pubmed.ncbi.nlm.nih.gov/23820514/

33. Damarell RA, Morgan DD, Tieman JJ. General practitioner strategies for managing patients with multimorbidity: A systematic review and thematic synthesis of qualitative research. BMC Fam Pract [Internet]. 2020 Jul 1 [cited 2023 May 25];21(1):1–23. Available from: https://bmcprimcare.biomedcentral.com/articles/10.1186/s12875-020-01197-8

34. Smith SM, Wallace E, Clyne B, Boland F, Fortin M. Interventions for improving outcomes in patients with multimorbidity in primary care and community setting: a systematic review. Syst Rev [Internet]. 2021 Dec 1 [cited 2023 May 25];10(1):1–23. Available from: https://systematicreviewsjournal.biomedcentral.com/articles/10.1186/s13643-021-01817-z

35. Excess winter mortality in England and Wales - Office for National Statistics [Internet]. [cited 2023 May 25]. Available from: https://www.ons.gov.uk/peoplepopulationandcommunity/birthsdeathsandmarriages/deaths/bulletins/excesswintermortalityinenglandandwales/2017to2018provisionaland2016to2017final

36. Kabir A, Tran A, Ansari S, Conway DP, Barr M. Impact of multimorbidity and complex multimorbidity on mortality among older Australians aged 45 years and over: a large population-based record linkage study. BMJ Open [Internet]. 2022 Jul 1 [cited 2023 May 25];12(7):e060001. Available from: https://bmjopen.bmj.com/content/12/7/e060001

37. Willadsen TG, Siersma V, Nicolaisdóttir DR, Køster-Rasmussen R, Jarbøl DE, Reventlow S, et al. Multimorbidity and mortality: A 15-year longitudinal registry-based nationwide Danish population study. J Comorbidity [Internet]. 2018 [cited 2023 May 25];8(1):2235042X18804063. Available from: /pmc/articles/PMC6194940/

38. Fleming DM, Taylor RJ, Haguinet F, Schuck-Paim C, Logie J, Webb DJ, et al. Influenza-attributable burden in United Kingdom primary care. Epidemiol Infect [Internet]. 2016 Feb 1 [cited 2023 May 25];144(3):537. Available from: /pmc/articles/PMC4714299/

39. Haug N, Deischinger C, Gyimesi M, Kautzky-Willer A, Thurner S, Klimek P. High-risk multimorbidity patterns on the road to cardiovascular mortality. BMC Med [Internet]. 2020 Mar 10 [cited 2023 May 25];18(1):1–12. Available from: https://bmcmedicine.biomedcentral.com/articles/10.1186/s12916-020-1508-1

40. Jani BD, Hanlon P, Nicholl BI, McQueenie R, Gallacher KI, Lee D, et al. Relationship between multimorbidity, demographic factors and mortality: findings from the UK Biobank cohort. BMC Med [Internet]. 2019 Apr 10 [cited 2023 May 25];17(1). Available from: /pmc/articles/PMC6456941/

41. de Boer IH, Khunti K, Sadusky T, Tuttle KR, Neumiller JJ, Rhee CM, et al. Diabetes Management in Chronic Kidney Disease: A Consensus Report by the American Diabetes Association (ADA) and Kidney Disease: Improving Global Outcomes (KDIGO). Diabetes Care [Internet]. 2022 Dec 1 [cited 2023 May 25];45(12):3075–90. Available from: https://diabetesjournals.org/care/article/45/12/3075/147614/Diabetes-Management-in-Chronic-Kidney-Disease-A

42. Multimorbidity: a priority for global health research. 2018;

43. Tran VT, Diard E, Ravaud P. Priorities to improve the care for chronic conditions and multimorbidity: a survey of patients and stakeholders nested within the ComPaRe e-cohort. BMJ Qual Saf [Internet]. 2021 Jul 1 [cited 2023 May 25];30(7):577–87. Available from: https://qualitysafety.bmj.com/content/30/7/577

44. Veronese N, Koyanagi A, Dominguez LJ, Maggi S, Soysal P, Bolzetta F, et al. Multimorbidity increases the risk of dementia: a 15 year follow-up of the SHARE study. Age Ageing. 2023 Apr 1;52(4).

45. Tinetti ME, Fried TR, Boyd CM. Designing health care for the most common chronic condition--multimorbidity. JAMA [Internet]. 2012 Jun 13 [cited 2023 May 25];307(23):2493–4. Available from: https://pubmed.ncbi.nlm.nih.gov/22797447/

46. Bodenheimer T, Wagner EH, Grumbach K. Improving primary care for patients with chronic illness. JAMA [Internet]. 2002 Oct 9 [cited 2023 May 25];288(14):1775–9. Available from: https://pubmed.ncbi.nlm.nih.gov/12365965/

47. Sullivan MK, Jani BD, McConnachie A, Hanlon P, McLoone P, Nicholl BI, et al. Hospitalisation events in people with chronic kidney disease as a component of multimorbidity: parallel cohort studies in research and routine care settings. BMC Med [Internet]. 2021 Dec 1 [cited 2023 May 25];19(1). Available from: /pmc/articles/PMC8603496/

48. Hay SI, Abajobir AA, Abate KH, Abbafati C, Abbas KM, Abd-Allah F, et al. Global, regional, and national disability-adjusted life-years (DALYs) for 333 diseases and injuries and healthy life expectancy (HALE) for 195 countries and territories, 1990-2016: A systematic analysis for the Global Burden of Disease Study 2016. Lancet. 2017 Sep 16;390(10100):1260–344.

49. Calvin CM, Conroy MC, Moore SF, Kuźma E, Littlejohns TJ. Association of Multimorbidity, Disease Clusters, and Modification by Genetic Factors With Risk of Dementia. JAMA Netw Open [Internet]. 2022 Sep 1 [cited 2023 May 25];5(9):e2232124–e2232124. Available from: https://jamanetwork.com/journals/jamanetworkopen/fullarticle/2796543

50. Diernberger K, Luta X, Bowden J, Droney J, Lemmon E, Tramonti G, et al. Variation in hospital cost trajectories at the end of life by age, multimorbidity and cancer type. Int J Popul data Sci [Internet]. 2023 [cited 2023 May 25];8(1):1768. Available from: https://pubmed.ncbi.nlm.nih.gov/36721848/

51. Muth C, Blom JW, Smith SM, Johnell K, Gonzalez-Gonzalez AI, Nguyen TS, et al. Evidence supporting the best clinical management of patients with multimorbidity and polypharmacy: a systematic guideline review and expert consensus. J Intern Med [Internet]. 2019 Mar 1 [cited 2023 May 25];285(3):272–88. Available from: https://onlinelibrary.wiley.com/doi/full/10.1111/joim.12842

52. Salisbury C, Man MS, Bower P, Guthrie B, Chaplin K, Gaunt DM, et al. Management of multimorbidity using a patient-centred care model: a pragmatic cluster-randomised trial of the 3D approach. Lancet (London, England) [Internet]. 2018 Jul 7 [cited 2023 May 25];392(10141):41. Available from: /pmc/articles/PMC6041724/

53. Michielsen L, Bischoff EWMA, Schermer T, Laurant M. Primary healthcare competencies needed in the management of person-centred integrated care for chronic illness and multimorbidity: Results of a scoping review. BMC Prim Care 2023 241 [Internet]. 2023 Apr 12 [cited 2023 May 25];24(1):1–13. Available from: https://bmcprimcare.biomedcentral.com/articles/10.1186/s12875-023-02050-4

54. Struckmann V, Leijten FRM, van Ginneken E, Kraus M, Reiss M, Spranger A, et al. Relevant models and elements of integrated care for multi-morbidity: Results of a scoping review. Health Policy (New York). 2018 Jan 1;122(1):23–35.

55. Boult C, Karm L, Groves C. Improving Chronic Care: The “Guided Care” Model. Perm J [Internet]. 2008 Mar [cited 2023 May 25];12(1):50. Available from: /pmc/articles/PMC3042340/

56. Blane DN, Lewandowska M. Living with cancer and multimorbidity: The role of primary care. Curr Opin Support Palliat Care [Internet]. 2019 Sep 1 [cited 2023 May 25];13(3):213–9. Available from: https://journals.lww.com/co-supportiveandpalliativecare/Fulltext/2019/09000/Living_with_cancer_and_multimorbiditythe_role_of.12.aspx

57. Ahmad T, Gopal D, Dayem Ullah AZM, Taylor S. Multimorbidity in patients living with and beyond cancer: protocol for a scoping review. BMJ Open [Internet]. 2022 May 1 [cited 2023 May 25];12(5):e057148. Available from: https://bmjopen.bmj.com/content/12/5/e057148

58. MacRae C, McMinn M, Mercer SW, Henderson D, McAllister DA, Ho I, et al. The impact of varying the number and selection of conditions on estimated multimorbidity prevalence: A cross-sectional study using a large, primary care population dataset. PLOS Med [Internet]. 2023 Apr 1 [cited 2023 Jun 1];20(4):e1004208. Available from: https://journals.plos.org/plosmedicine/articleid=10.1371/journal.pmed.1004208

